# Protocol for DNA Extraction from QuantiFERON-TB Gold Tubes for PCR and Sequencing Applications

**DOI:** 10.64898/2026.03.16.26348529

**Authors:** Urooj Subhan, Zaineb Akram, Sara Shafqat, Sidra Younis

**Author notes:** **Corresponding author:** Dr. Sidra Younis, PhD, Associate Professor, Department of Biological Sciences, National University of Medical Sciences, Rawalpindi, Pakistan.

## Abstract

Latent tuberculosis infection (LTBI) remains a significant barrier to global TB control and elimination efforts. The QuantiFERON-TB Gold (QFT) assay is commonly used for the diagnosis of LTBI. However, blood collected in QFT tubes is seldom utilized for molecular and genetic analysis due to the presence of heparin and a dense gel barrier that hinders efficient DNA extraction. To address this limitation, we aimed to develop a method for directly isolating high-quality DNA from blood in QFT tubes, eliminating the need for additional blood sampling and enabling their use in both diagnostic and molecular workflows.

In this study, DNA was extracted from blood in EDTA and QFT tubes using a hybrid approach that combined manual lysis with three commercial kits: Thermo Scientific GeneJET, QIAamp DNA Blood Kit, and FavorPrep™ Blood Genomic DNA Extraction Kit. DNA concentration and purity were measured with a Multiskan SkyHigh Microplate Spectrophotometer, while integrity was assessed through agarose gel electrophoresis. Two nucleic acid amplification techniques (NAATs), ARMS-PCR and whole exome sequencing (WES) were performed to validate applicability of extracted DNA for molecular biology applications.

We did not find any differences in the quantity, quality, or application of PCR or sequencing for DNA extracted from EDTA or QFT tubes. The extracted DNA from both EDTA and QFT tubes exhibited A260/280 ratios of 1.7-1.9 and concentrations ranging from 4.9 to 118.5 µg/mL, indicating an adequate yield and purity. Intact genomic DNA and PCR product bands on agarose gel indicated suitability for downstream applications. Additionally, WES produced 6.47-8.71 GB of data per sample, with 42.8-57.7 M reads and GC content between 49.29% and 52.54%. Sequencing metrics were consistently strong, with Q20 values exceeding 98.6% and Q30 values above 95%.

Our study presents an optimized and reproducible protocol for extracting high-quality DNA from QFT tubes, producing DNA suitable for both PCR and sequencing technologies. This protocol provides a cost-effective and practical strategy to integrate LTBI diagnosis with genomic research, particularly beneficial in resource-limited settings. This study introduces a novel analytical workflow applicable to diagnostic laboratory settings, enabling the integration of routine LTBI immunodiagnostic testing with downstream genomic analysis. The approach supports improved utilization of clinical specimens in laboratory medicine and may facilitate future biomarker and precision diagnostics research.

## Introduction

LTBI remains a major barrier to global tuberculosis (TB) elimination programs, with nearly one-quarter of the world’s population estimated to harbor *Mycobacterium tuberculosis* (*Mtb*) in a dormant state (Kiazyk & Ball, 2017). Although individuals with LTBI are asymptomatic, they still carry a lifelong risk of reactivation (particularly under conditions of immunosuppression) (Mohammadnabi et al., 2024). Early identification of LTBI is crucial, as it enables timely preventive treatment to reduce the progression to active TB (ATB), helps to control the spread of disease, and supports targeted preventive therapy (Gunasekaran, Ranganathan, & Bethunaickan, 2025; Miggiano, Rizzi, & Ferraris, 2020).

The QFT assay, an interferon-gamma release assay (IGRA), is a widely employed assay for the detection of LTBI. This diagnostic tool measures interferon-gamma (IFN-γ) release from T-cells upon stimulation with *Mtb*-specific antigens (ESAT-6 and CFP-10). Despite its diagnostic value, the QFT assay presents several limitations, especially in resource-limited settings (Sayin, Alci, Ozanat, Duman, & Karahasan, 2025; Sester et al., 2025). In Pakistan, the high cost of the test and the requirement of specialized tubes for its performance restrict routine use, with many laboratories conducting the assay only weekly or monthly. Consequently, access to LTBI diagnosis remains limited for the potentially infected population. Nucleic acid amplification tests (NAATs) have potential for the identification of sensitive diagnostic procedures. Molecular and genetic studies on LTBI patients require reliable sources of DNA. However, collecting parallel samples in EDTA tubes for nucleic acid extraction alongside QFT tubes for diagnosis is not feasible in most clinical and research contexts. LTBI status is confirmed only after QFT assay results are available, making it impractical to collect additional EDTA samples retrospectively. Furthermore, patients are often unwilling to undergo repeated venipuncture, while multiple blood draws increase both logistical burden and study costs. These barriers pose significant challenges for researchers seeking to conduct large-scale DNA-based studies on LTBI populations.

QFT tubes, if optimized for nucleic acid recovery, could serve as a valuable dual-purpose resource. However, their utility for DNA extraction has been limited by technical constraints. Lithium heparin present in QFT tubes inhibits PCR and sequencing, while the dense gel barrier complicates recovery of cellular material. The gel’s sticky nature often clogs pipette tips and interferes with cellular pellet handling, making extraction labor-intensive and inefficient. To date, no standardized protocol has been reported for efficient DNA extraction from QFT tubes.

In this study, we developed and optimized a protocol for extracting high-quality genomic DNA directly from QFT tubes. DNA was extracted from both EDTA and QFT tubes to compare yield, purity, and downstream performance in PCR and sequencing. Three commercially available kits with modified extraction strategies were employed and evaluated for the extraction of the best quality DNA, and the protocol was refined to overcome the inhibitory and technical challenges inherent to QFT samples.

Our optimized protocol demonstrates that QFT tubes can serve as a reliable and previously underutilized source of genomic DNA for downstream molecular analyses. By enabling direct DNA extraction from routinely collected diagnostic specimens, this workflow eliminates the need for additional blood sampling while providing a cost- and time-efficient solution within clinical laboratory practice. From a laboratory medicine perspective, maximizing the analytical value of diagnostic samples is essential for integrating immunodiagnostic testing with emerging molecular and genomic technologies. The proposed approach therefore represents a practical methodological advancement that enhances sample utilization in routine diagnostic workflows and supports future biomarker discovery, large-scale epidemiological investigations, and data-driven diagnostic strategies, particularly in resource-limited healthcare settings.

## Material and Methods

### Settings

The study received ethical approval from the Institutional Review Board Committee of the National University of Medical Sciences, Islamabad, in accordance with the Declaration of Helsinki. Written informed consent was obtained from all participants before sample collection. Blood samples were collected from 15 individuals at Holy Family Hospital, Islamabad. Of these, 10 samples were drawn into QFT tubes, while the remaining 5 samples were collected in EDTA tubes for comparative analysis.

### DNA extraction

DNA was extracted from the EDTA tubes directly by following standard kits including Thermo Scientific GeneJET, QIAamp DNA Blood Kit, and FavorPrep™ Blood Genomic DNA Extraction protocol. For QFT tubes, a modified procedure was applied.

1. After collecting and storing the serum, each of the QFT tubes was centrifuged in an inverted position for 5 min at 4400 rpm, which allowed the gel to move upward and the blood cells to concentrate near the cap. Clotted cells were transferred from each QFT tube to 15 mL falcon tube containing RBCs Lysis Buffer (0.32 M Sucrose, 10 mM Tris, and 5 mM MgCl_2_) and pipetted up and down several time until pipette tip is cleared and small about of this lysis buffer was transferred to the cap of each QFT tubes to dissolve and collect the cells stacked to the cap. A small amount of buffer is added to the QFT tube to dissolve the remaining cells stuck to the gel, and then transferred carefully to the RBCs lysis buffer. Precautions should be taken to avoid picking up the gel.
2. The tubes were then vortexed thoroughly to disrupt clots in the RBC lysis buffer and incubated at -20 °C for 20 min with vortexing occasionally. This was followed by centrifugation at 4400 rpm for 30 min. If the pellet was not sufficient, centrifugation was repeated for an additional 10 min. The supernatant was discarded, and the pellet was resuspended in 200 µL PBS, vortexed until fully dissolved, and centrifuged briefly for 10 s. The resuspended cells in PBS were then transferred into a freshly labeled Eppendorf tube.
3. 20 µL Proteinase K was added to the sample and mixed by pipetting up and down several times. This was followed by the addition of 200 µL FABG buffer, and the mixture was pulse-vortexed to ensure thorough mixing (don’t add Proteinase K directly to the buffer). The samples were incubated at 60 °C for 15 min with vortexing every 3–5 min, and tubes were briefly spun to collect drops inside the lid.
4. Next, 200 µL of 100% ethanol was added to the sample, mixed by pulse vortexing, and centrifuged briefly. The entire mixture, including any precipitate, was transferred into a FABG Mini Column placed in a collection tube and incubated for 5 min. Columns were centrifuged at maximum speed (17,000 × g) for 2 min, then placed into fresh collection tubes. A total of 400 µL W1 buffer was added, and the column was centrifuged at maximum speed (17,000 × g) for 1 min; the flow-through was discarded. This was followed by 750 µL Wash Buffer, centrifuged at maximum speed (17,000 × g) for 1 min, and the flow-through discarded. The column was further centrifuged at maximum speed for 3 min to ensure complete drying, a critical step to prevent residual ethanol from interfering with subsequent enzymatic reactions.
5. For elution, the FABG Mini Column was placed into a clean elution tube. A volume of 50 µL pre-warmed Elution Buffer or nuclease-free water (pH 7.5–9.0) was applied directly to the membrane center, ensuring complete absorption. The column was left to stand for 5 minutes and then centrifuged at maximum speed for 3 min to recover DNA. The eluted DNA was incubated at 55 °C for 5 min to facilitate complete dissolution and mixed gently by low-speed vortexing. DNA concentration and purity were assessed by the Multiskan SkyHigh Microplate Spectrophotometer and 1% agarose gel electrophoresis.
6. If the concentration and purity were satisfactory, DNA was stored at 4 °C. In cases where the yield was insufficient, re-elution was performed by loading the previously extracted DNA back into the column and repeating the elution steps (Step 5). For longterm storage, samples were preserved at -20 °C.
7. Precautionary note: The gel present in QFT tubes was extremely sticky and often adhered to pipette tips, making it difficult to recover blood cells without contamination. This also complicated the collection of intact pellets, requiring careful handling to avoid loss of material during processing.

### PCR Procedure

To confirm that DNA extracted from QFT tubes using a modified protocol is equally suitable for PCR, we performed PCR with previously designed primers and an optimized protocol from an earlier laboratory study (Subhan et al., 2026). The PCR procedure is detailed below. Primers were designed using Primer 1 software. The tetra-arms PCR using two sets of primers was used to selectively amplify the target region of interest. Gradient PCR was used for the optimization of the annealing temperature. PCR products were separated on 2% agarose gel and visualized by the Gel Doc XR+ Gel Documentation System (Bio-Rad). Nucleotide sequences of primers with their properties and PCR cycle details are given in Tables 1 and 2.

**Table 1:**
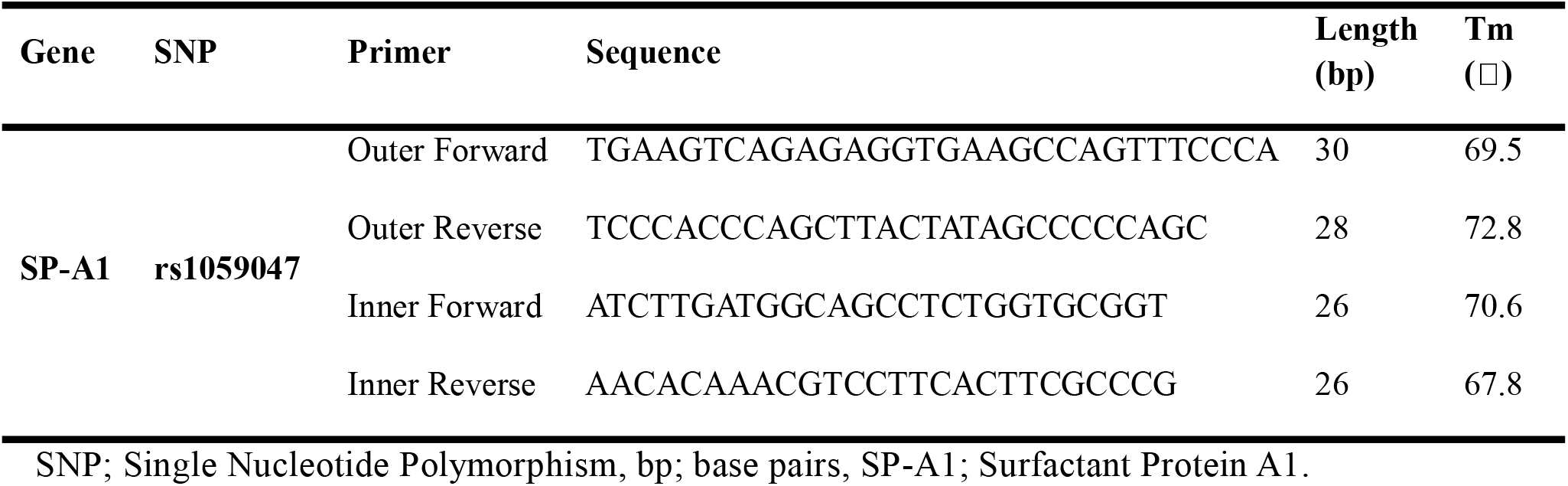
Sequence and properties of primers used for PCR.

**Table 2:** PCR conditions used for ARMS-PCR.

| SNP | PCR cycle details |  |  |  |  |
| --- | --- | --- | --- | --- | --- |
|  | Initial denaturation | Denaturation | Annealing<br>(35 cycles) | Extension | Final extension |
| rs1059047 | 95 °C for 5 min | 95 °C for 1 min | 69 °C for 1 min | 72 °C for 1 min | 72 °C for 10 min |

**Table 3:**
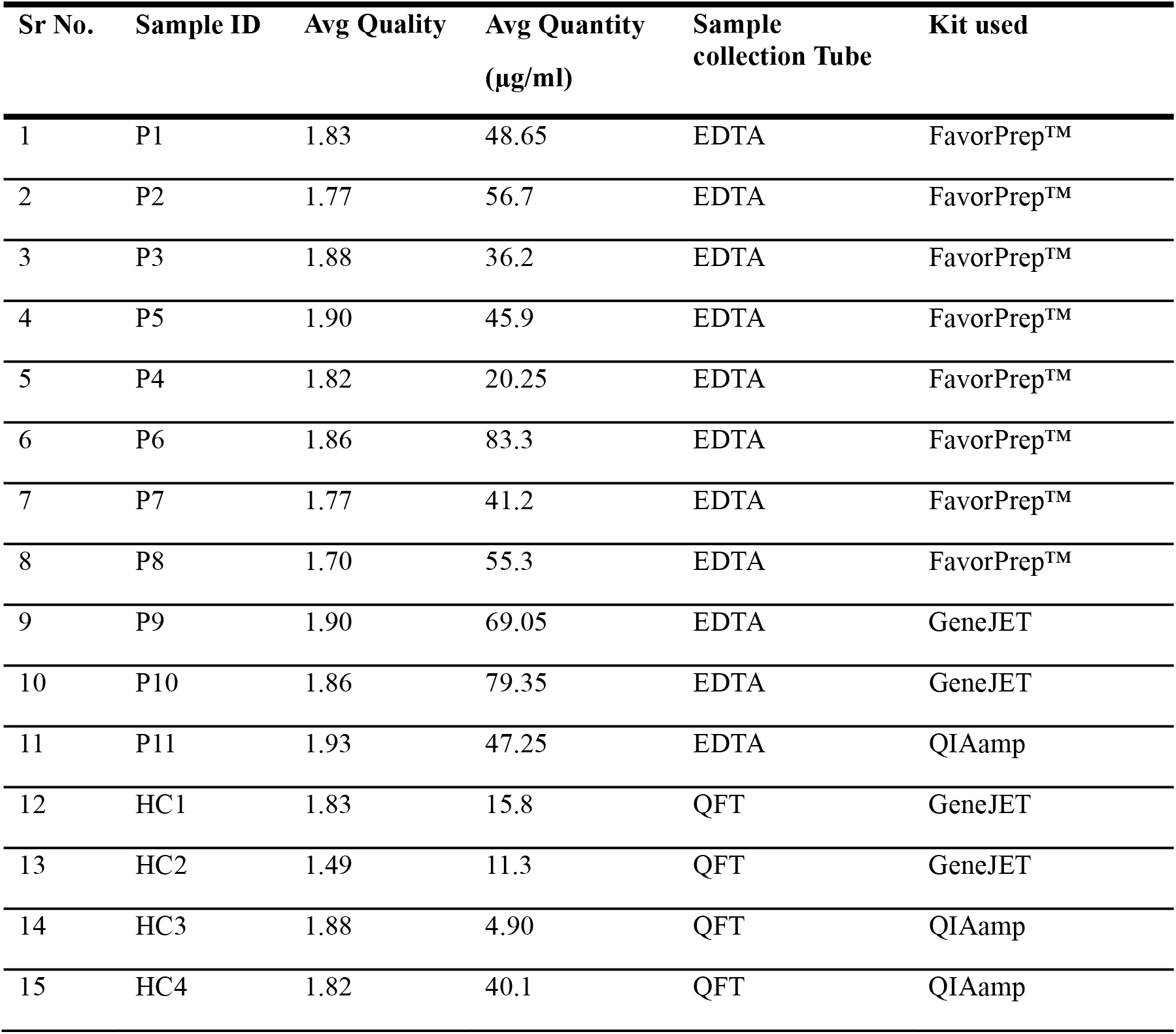
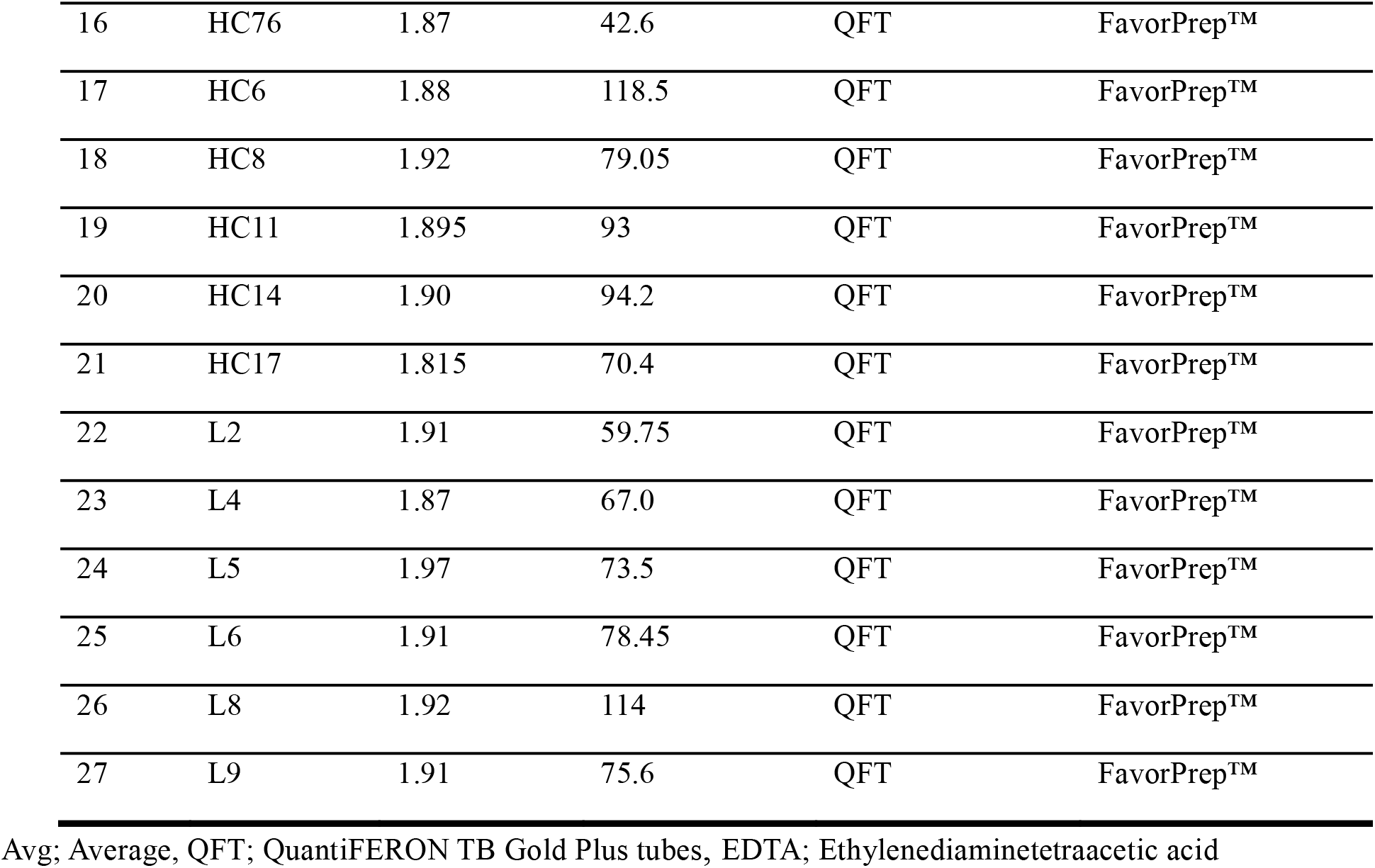
Quality analysis of genomic DNA by UV absorption at 260 and 280 nm.

| Sr No. | Sample ID | Avg Quality | Avg Quantity<br>(µg/ml) | Sample<br>collection Tube | Kit used |
| --- | --- | --- | --- | --- | --- |
| 1 | P1 | 1.83 | 48.65 | EDTA | FavorPrep™ |
| 2 | P2 | 1.77 | 56.7 | EDTA | FavorPrep™ |
| 3 | P3 | 1.88 | 36.2 | EDTA | FavorPrep™ |
| 4 | P5 | 1.90 | 45.9 | EDTA | FavorPrep™ |
| 5 | P4 | 1.82 | 20.25 | EDTA | FavorPrep™ |
| 6 | P6 | 1.86 | 83.3 | EDTA | FavorPrep™ |
| 7 | P7 | 1.77 | 41.2 | EDTA | FavorPrep™ |
| 8 | P8 | 1.70 | 55.3 | EDTA | FavorPrep™ |
| 9 | P9 | 1.90 | 69.05 | EDTA | GeneJET |
| 10 | P10 | 1.86 | 79.35 | EDTA | GeneJET |
| 11 | P11 | 1.93 | 47.25 | EDTA | QIAamp |
| 12 | HC1 | 1.83 | 15.8 | QFT | GeneJET |
| 13 | HC2 | 1.49 | 11.3 | QFT | GeneJET |
| 14 | HC3 | 1.88 | 4.90 | QFT | QIAamp |
| 15 | HC4 | 1.82 | 40.1 | QFT | QIAamp |
| 16 | HC76 | 1.87 | 42.6 | QFT | FavorPrep™ |
| 17 | HC6 | 1.88 | 118.5 | QFT | FavorPrep™ |
| 18 | HC8 | 1.92 | 79.05 | QFT | FavorPrep™ |
| 19 | HC11 | 1.895 | 93 | QFT | FavorPrep™ |
| 20 | HC14 | 1.90 | 94.2 | QFT | FavorPrep™ |
| 21 | HC17 | 1.815 | 70.4 | QFT | FavorPrep™ |
| 22 | L2 | 1.91 | 59.75 | QFT | FavorPrep™ |
| 23 | L4 | 1.87 | 67.0 | QFT | FavorPrep™ |
| 24 | L5 | 1.97 | 73.5 | QFT | FavorPrep™ |
| 25 | L6 | 1.91 | 78.45 | QFT | FavorPrep™ |
| 26 | L8 | 1.92 | 114 | QFT | FavorPrep™ |
| 27 | L9 | 1.91 | 75.6 | QFT | FavorPrep™ |

Avg; Average, QFT; QuantiFERON TB Gold Plus tubes, EDTA; Ethylenediaminetetraacetic acid

**Table 4:**
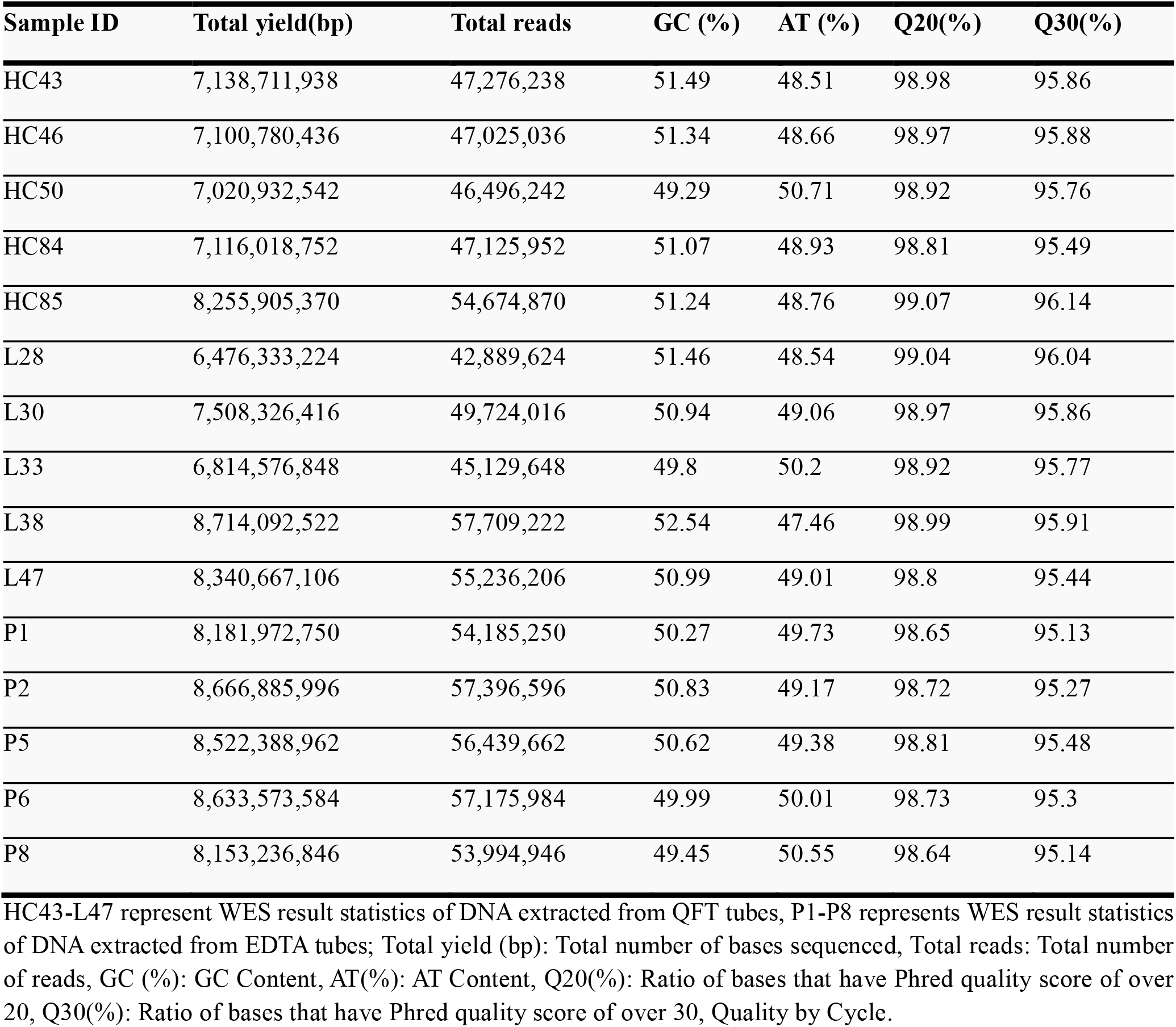
Fastq statistics of DNA samples extracted from EDTA and QFT tubes.

| Sample ID | Total yield(bp) | Total reads | GC (%) | AT (%) | Q20(%) | Q30(%) |
| --- | --- | --- | --- | --- | --- | --- |
| HC43 | 7,138,711,938 | 47,276,238 | 51.49 | 48.51 | 98.98 | 95.86 |
| HC46 | 7,100,780,436 | 47,025,036 | 51.34 | 48.66 | 98.97 | 95.88 |
| HC50 | 7,020,932,542 | 46,496,242 | 49.29 | 50.71 | 98.92 | 95.76 |
| HC84 | 7,116,018,752 | 47,125,952 | 51.07 | 48.93 | 98.81 | 95.49 |
| HC85 | 8,255,905,370 | 54,674,870 | 51.24 | 48.76 | 99.07 | 96.14 |
| L28 | 6,476,333,224 | 42,889,624 | 51.46 | 48.54 | 99.04 | 96.04 |
| L30 | 7,508,326,416 | 49,724,016 | 50.94 | 49.06 | 98.97 | 95.86 |
| L33 | 6,814,576,848 | 45,129,648 | 49.8 | 50.2 | 98.92 | 95.77 |
| L38 | 8,714,092,522 | 57,709,222 | 52.54 | 47.46 | 98.99 | 95.91 |
| L47 | 8,340,667,106 | 55,236,206 | 50.99 | 49.01 | 98.8 | 95.44 |
| P1 | 8,181,972,750 | 54,185,250 | 50.27 | 49.73 | 98.65 | 95.13 |
| P2 | 8,666,885,996 | 57,396,596 | 50.83 | 49.17 | 98.72 | 95.27 |
| P5 | 8,522,388,962 | 56,439,662 | 50.62 | 49.38 | 98.81 | 95.48 |
| P6 | 8,633,573,584 | 57,175,984 | 49.99 | 50.01 | 98.73 | 95.3 |
| P8 | 8,153,236,846 | 53,994,946 | 49.45 | 50.55 | 98.64 | 95.14 |

HC43-L47 represent WES result statistics of DNA extracted from QFT tubes, P1-P8 represents WES result statistics of DNA extracted from EDTA tubes; Total yield (bp): Total number of bases sequenced, Total reads: Total number of reads, GC (%): GC Content, AT(%): AT Content, Q20(%): Ratio of bases that have Phred quality score of over 20, Q30(%): Ratio of bases that have Phred quality score of over 30, Quality by Cycle.

### Whole Exome sequencing

To assess whether the extracted DNA was suitable for whole exome sequencing (WES), we performed WES on DNA obtained from QFT tubes and, for comparison, on DNA extracted from EDTA tubes. A total of 15 samples including 10 in QFT tubes and 5 in EDTA tubes were subjected to WES. Library preparation was performed using the Twist Bioscience protocol, and sequencing was conducted on the Illumina platform (Korea Macrogen) with sequencing-by-synthesis chemistry.

## Results

### Nanodrop and Gel Electrophoresis

DNA extraction was carried out several times using manual method and different commercial kits including the Thermo Scientific GeneJET Genomic DNA Purification Kit, the QIAamp DNA Blood Kit, and the FavorPrep™ Blood Genomic DNA Extraction Kit. The concentration and purity of the extracted DNA were evaluated using a Multiskan SkyHigh Microplate Spectrophotometer (Bio-Rad) and further verified by 1% agarose gel electrophoresis (Supplementary File 1). Measurements were performed in duplicate, and the mean values for concentration and purity of each sample are presented in Table 1. The integrity of the extracted DNA, as visualized through gel electrophoresis, is shown in Figure 1.

**Figure 1:**
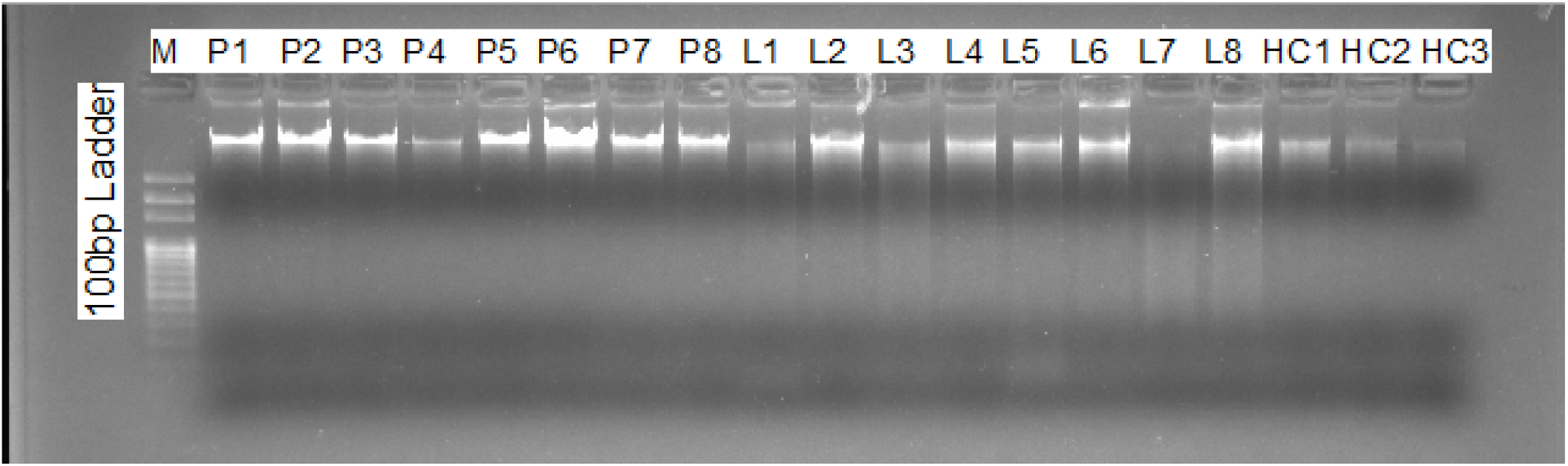
Representative Agarose gel picture showing extracted DNA. M represents 100 bp DNA ladder. P1 to P6 lanes represents the DNA’s extracted from EDTA tubes; L1 to HC3 lanes represent the DNA extracted from QFT tubes.

### PCR reaction for DNA extracted from EDTA and QFT tubes

ARMS-PCR was performed to evaluate suitability of DNA extracted from QFT tubes for molecular biology applications, with DNA extracted from EDTA tubes used for comparison. The PCR products were resolved on a 2% agarose gel, and the results are shown in Figure 2. Clear amplification bands confirmed that the extracted DNA using the modified protocol is perfectly suitable for downstream molecular biology applications.

**Figure 2:**
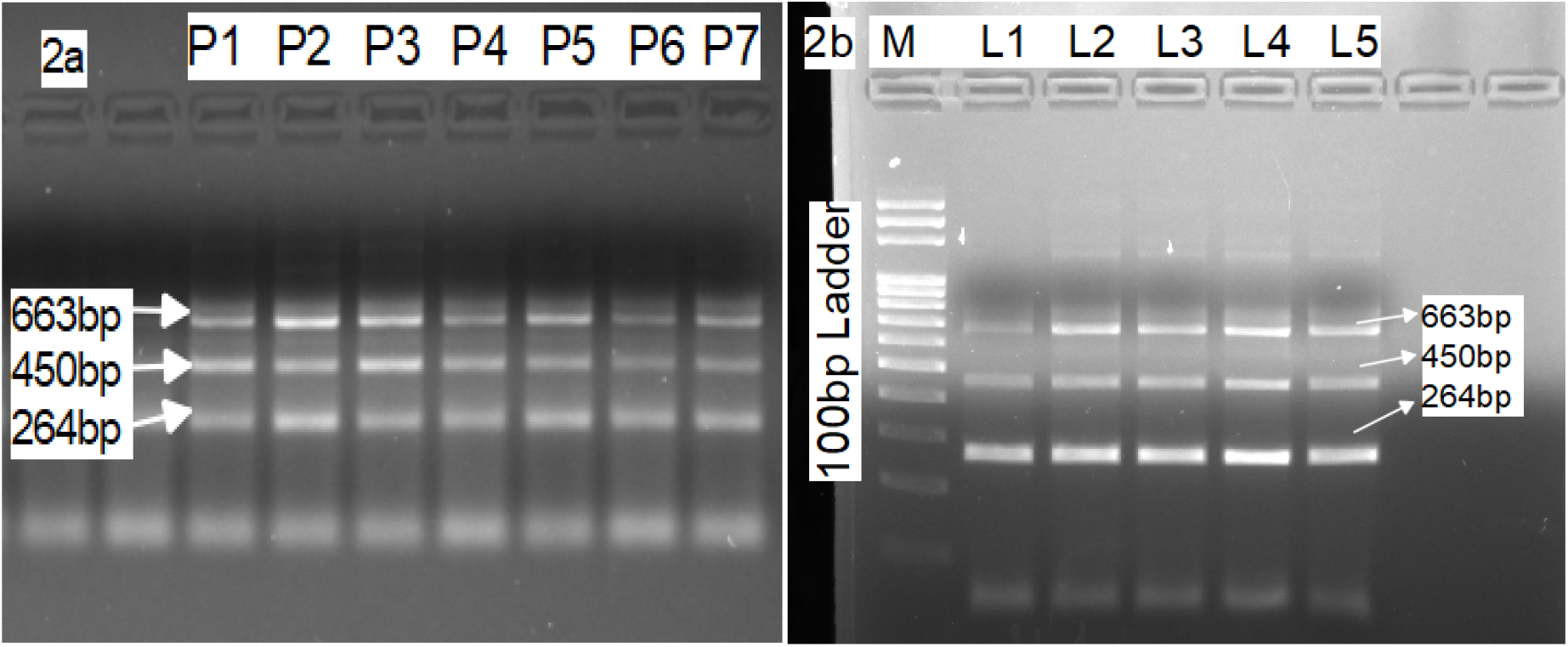
Agarose gel picture representing genotype pattern for Tetra-Arms PCR (rs1059047) for DNA extracted from EDTA and QFT tubes. Lane M represents 100 bp DNA ladder. Fig 2a: The P1 to P7 lanes represent ARMC-PCR for the DNA samples extracted from EDTA tubes. Fig 2b: The L1 to L5 lanes represent ARMC-PCR for the DNA samples extracted from QFT tubes. Product sizes are characterized as 663+450 bp for CC, 663+264 bp for TT, and 663+450+264 bp for CT.

**Figure 3:**
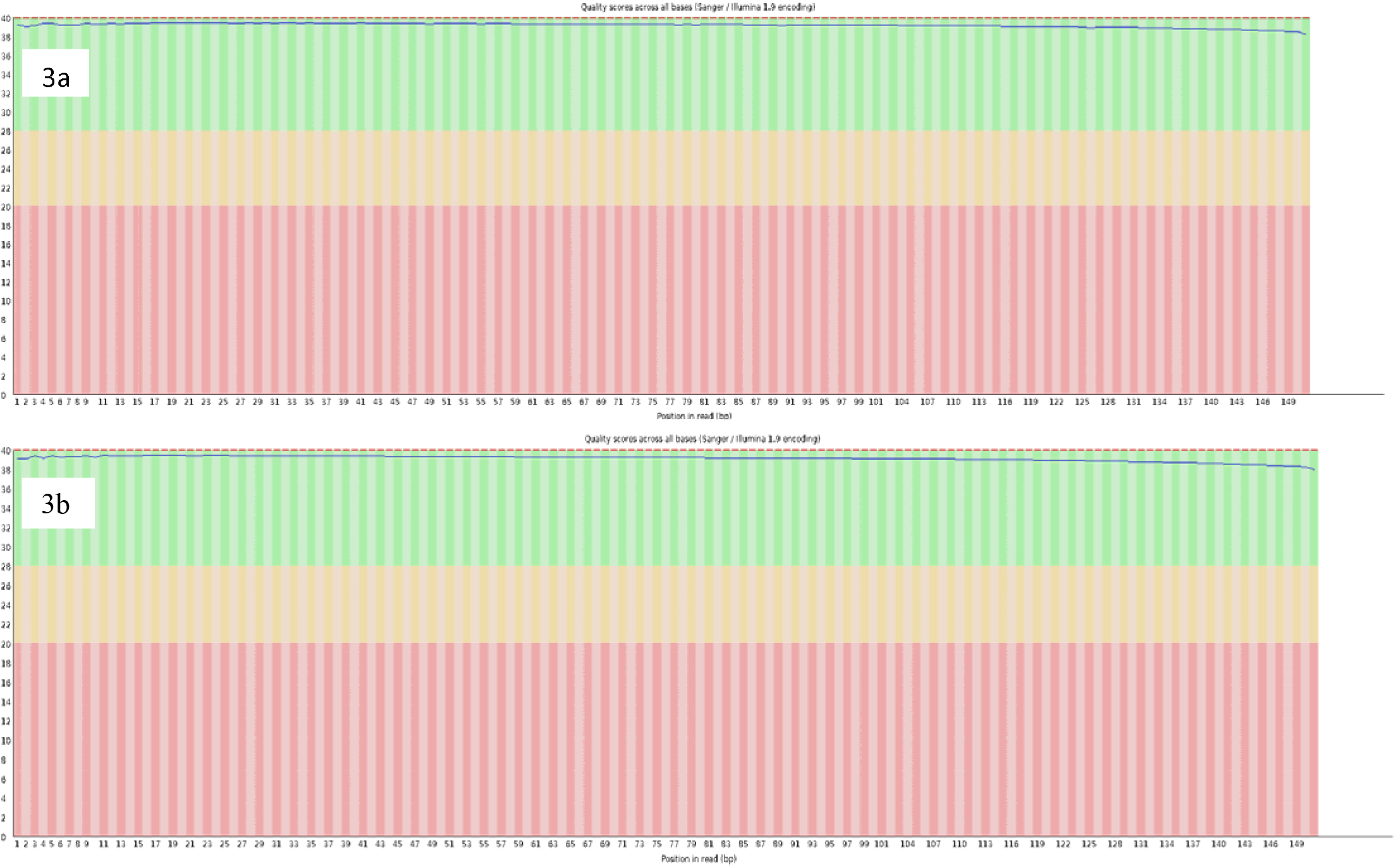
Quality scores of all bases. Representative quality control (QC) plots showing base quality scores across all bases. Fig 3a: QC of DNA extracted from QFT tubes and Fig 3b: QC of DNA extracted from EDTA tubes.

### WES for the DNA extracted from EDTA and QFT tubes

WES was successfully performed for all 15 DNA samples, including 10 samples extracted from the modified protocol and 5 DNA samples extracted from EDTA tubes. We obtained high-quality sequencing results for all of the 15 samples. The total sequencing yield ranged from 6.47 to 8.71 GB per sample, with the number of total reads varying between 42.8 and 57.7 M. The GC content across all samples was consistent, ranging from 49.29% to 52.54%, with corresponding AT content values between 47.46% and 50.71%, indicating balanced nucleotide representation.

The sequencing quality was high across all samples. The proportion of bases with a Phred quality score ≥ 20 (Q20) exceeded 98.7% in all cases, while the proportion of bases with a Phred score ≥ 30 (Q30) remained above 95.1%. These values confirm that the sequencing runs on DNA extracted from QFT tubes produced reliable and high-quality data suitable for downstream genomic analyses, including variant calling and comparative studies.

## Discussion

LTBI remains one of the major challenges in the global effort to eliminate TB. Individuals with LTBI harbor *Mtb* bacilli in a dormant state without clinical symptoms, yet they serve as a potential reservoir for future disease reactivation. The diagnosis of LTBI is difficult because of the absence of overt symptoms and the limitations of existing diagnostic tools. Among the available methods, the QFT assay is widely used for detecting *Mtb* infection through interferon-gamma release. However, samples collected in QFT tubes are primarily intended for immunological assays, and extracting high-quality genomic DNA from these tubes for molecular studies presents significant technical difficulties due to low cell counts, the presence of stabilizing agents, and RNA-preserving additives that can interfere with DNA isolation.

In this study, we focused on optimizing DNA extraction from QFT tubes for downstream genomic applications, including PCR and WES. Initially, DNA extraction was performed using several commercially available kits, including Thermo Scientific GeneJET Genomic DNA Purification Kit, QIAamp DNA Blood Kit, and FavorPrep™ Blood Genomic DNA Extraction Kit. Although both the Qiagen and Thermo Scientific kits yielded DNA of good purity and integrity, their high cost limits their large-scale application in resource-constrained settings. Therefore, to develop a cost-effective and efficient extraction approach, we evaluated the FavorPrep™ kit and further optimized it by combining it with a modified manual extraction step. This hybrid approach enhanced cell lysis and nucleic acid recovery, resulting in DNA yields exceeding 100 ng/µL (concentrations sufficient for next-generation sequencing (NGS) and other molecular assays).

Among the evaluated methods, the modified FavorPrep™ protocol yielded consistently higher DNA from both EDTA and QFT samples, supporting its application in large-scale molecular studies. DNA quality assessment using a Multiskan SkyHigh Microplate Spectrophotometer showed A260/A280 ratios of 1.7–1.9, while intact genomic DNA and clear PCR bands on 2% agarose gels confirmed preserved integrity and absence of inhibitors. WES of the extracted DNA produced consistently high-quality datasets, with per-sample yields ranging from 6.47 to 8.71 GB. GC content was uniform across all samples (49%–52%), and sequencing quality metrics were robust, as reflected by Phred scores exceeding 98% at Q20 and 95% at Q30. These parameters indicate efficient library preparation and reliable sequencing performance. The strong WES quality metrics observed in this study demonstrate that the DNA extracted using the modified protocol retained sufficient integrity and purity (Delahaye & Nicolas, 2021; So et al., 2018). Moreover, the stable and balanced GC content suggests uniform genome representation, which is critical for dependable variant calling and meaningful downstream comparative analysis.

Overall, the study successfully demonstrates that DNA of high molecular quality can be extracted directly from QFT tubes (a type of sample not originally designed for DNA work) by adopting a combined manual and kit-based approach. The modified FavorPrep™ method not only maintains DNA purity and integrity but also offers a cost-effective alternative to commercially expensive kits. This approach enables the integration of QFT-based immunodiagnostic sampling with genetic and genomic studies of TB and LTBI, facilitating molecular research without additional invasive sample collection.

## Conclusion

To best of our knowledge, this is first study to optimize and report a cost-effective protocol for DNA extraction from QFT tubes for PCR and sequencing applications. The optimized method yielded high-quality and adequate-quantity DNA suitable for molecular analyses. The integrity and purity of extracted DNA were confirmed not only through standard quality assessment methods but also through successful downstream applications including PCR and WES. These findings demonstrate that extracted DNA was free from significant contaminants and can be efficiently used in advanced molecular biology technologies.

Major strength of this study is development and validation of a novel, optimized protocol that expands utility of QFT tubes beyond routine IFN-γ testing. Overall, this work provides a practical foundation for utilizing QFT tubes in genetic and sequencing-based research and represents a useful analytical approach for improving specimen utilization within clinical laboratory diagnostics.

## Data Availability

All the data produced in the present study are available upon reasonable request to the authors.

## Acknowledgments

This study was funded by the National University of Medical Sciences (NUMS), Rawalpindi, Pakistan, and by the Higher Education Commission of Pakistan through the NRPU project (HEC-NRPU-15101).

## Availability of Data and Materials

All data generated or analyzed during this study are included in this published article [and its supplementary information files].

## Authors’ contributions

Urooj Subhan: Data Curation, Formal Analysis, Investigation, Writing Original Draft

Dr. Zaineb Akram: Visualization, Validation, Data Curation, Methodology,

Dr. Sidra Younis: Conceptualization, Project Administration, Methodology, Supervision, Visualization, Validation, Writing-Review, and Editing

Resources and Funding Acquisition: National University of Medical Sciences, Rawalpindi, and HEC-NRPU.

