## Supplementary File for "Protocol for DNA Extraction from QuantiFERON-TB Gold Tubes for PCR and Sequencing Applications"

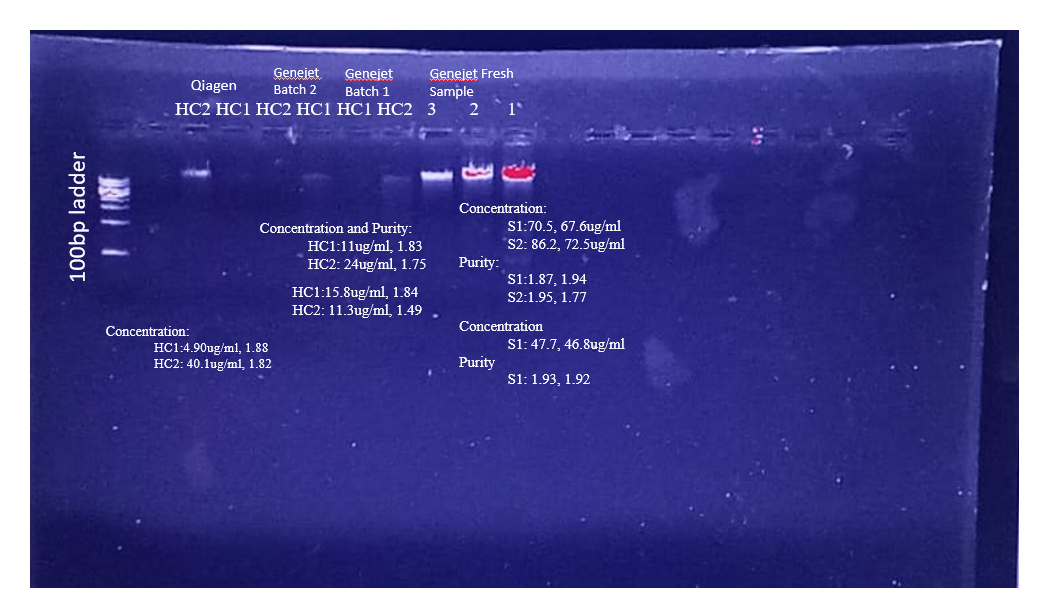
**S1: Comparison of concentration and purity of DNA extracted using different kits.**

**S2: Quality scores of all bases among all samples**

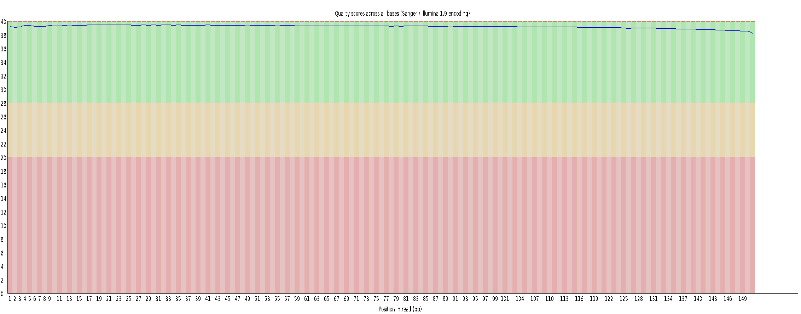

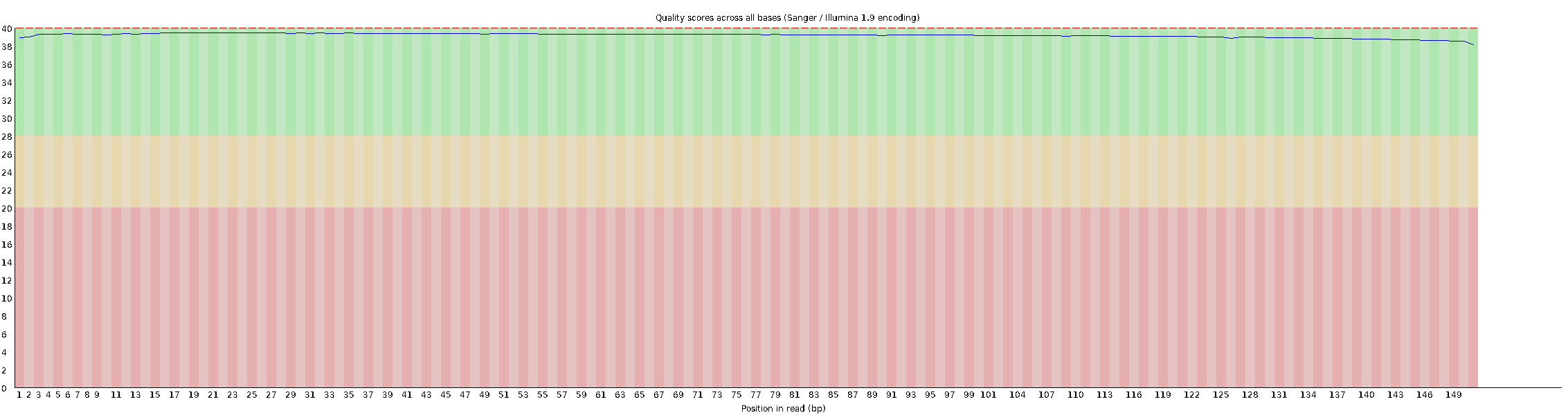

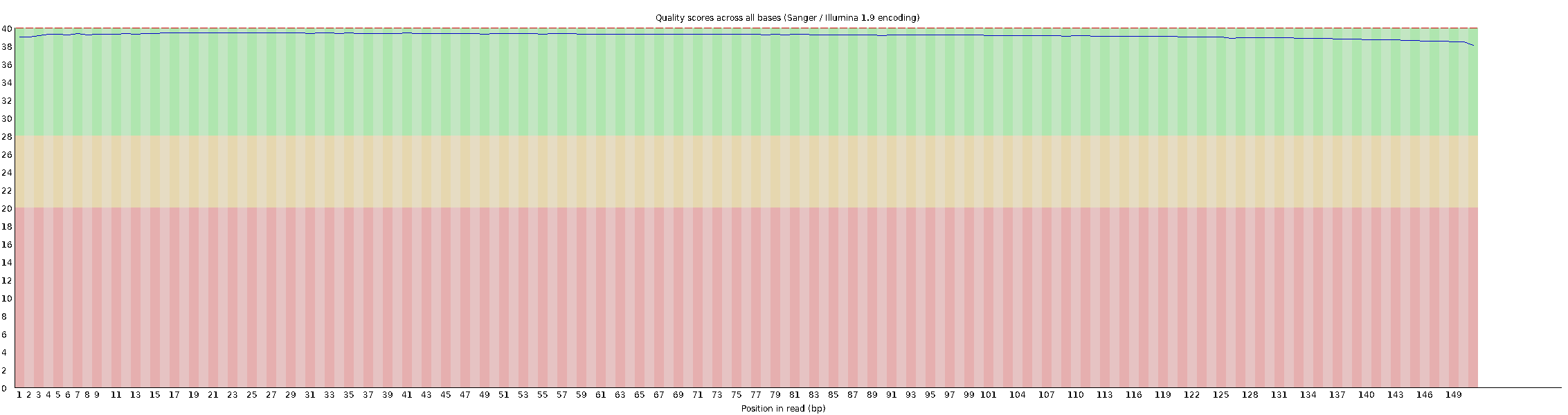

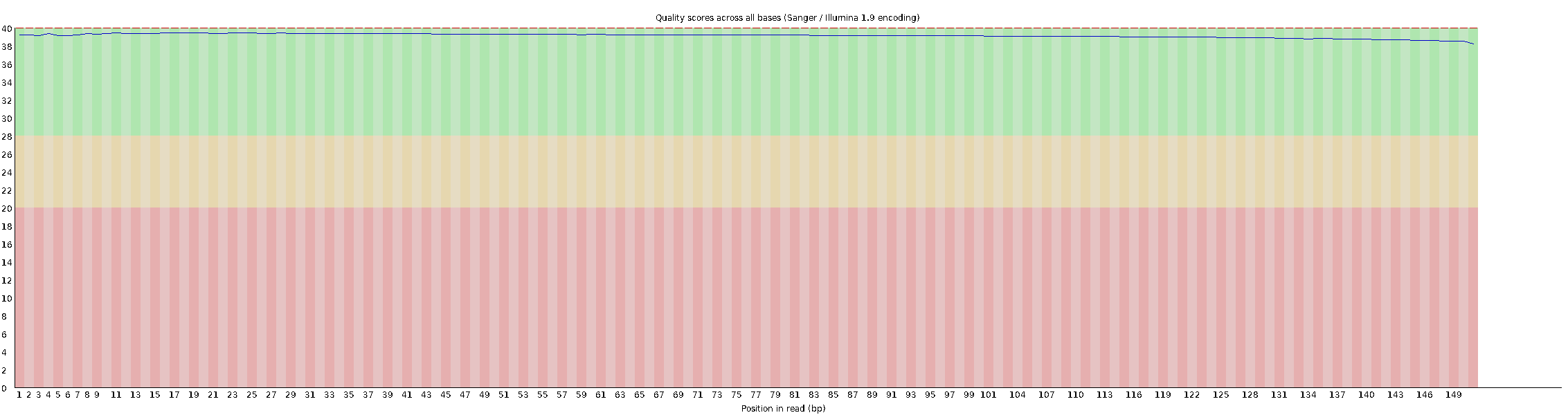

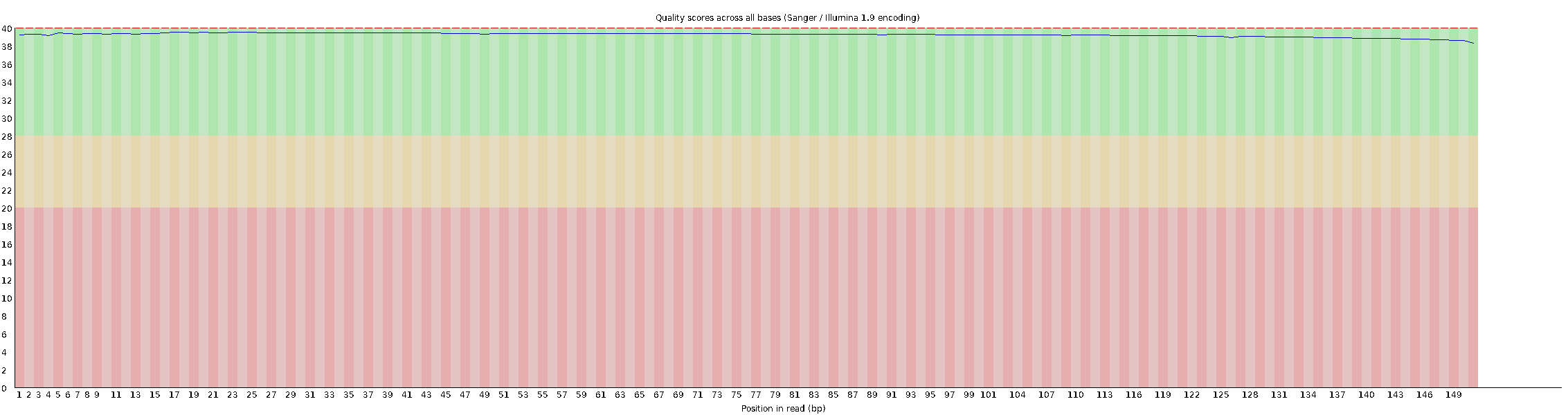

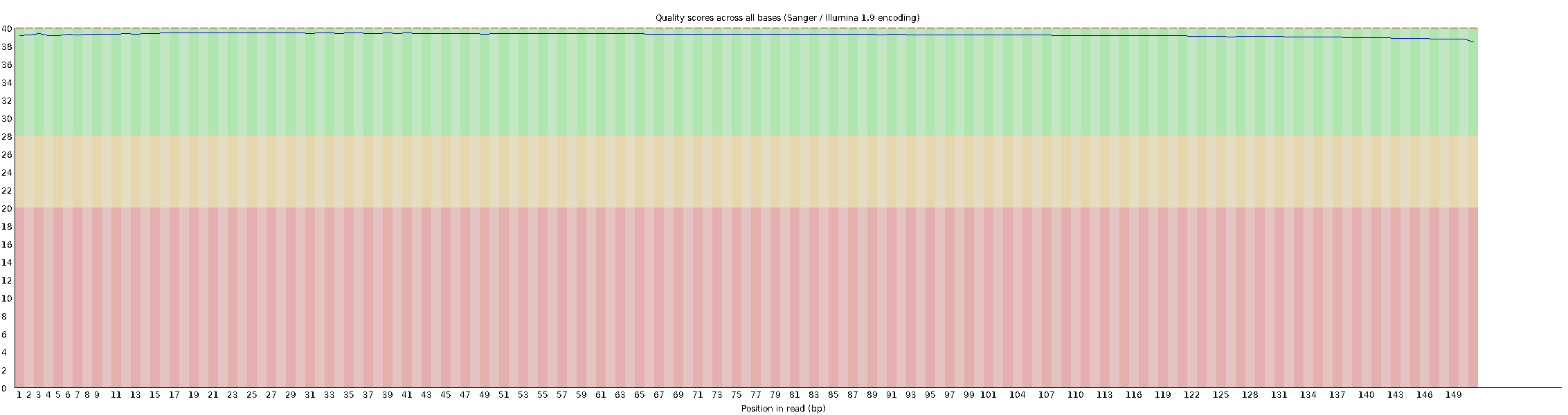

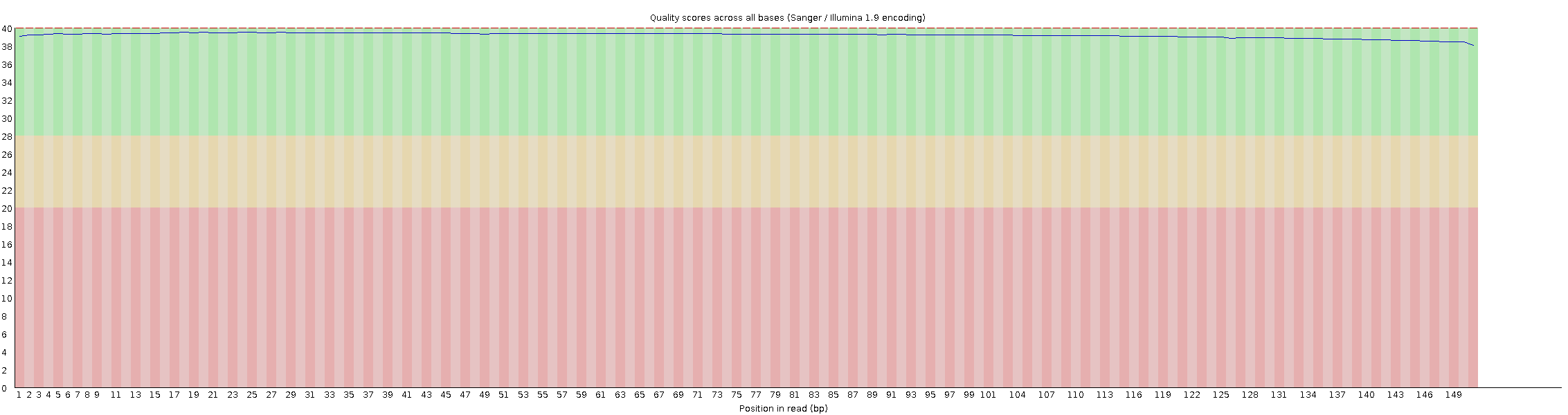

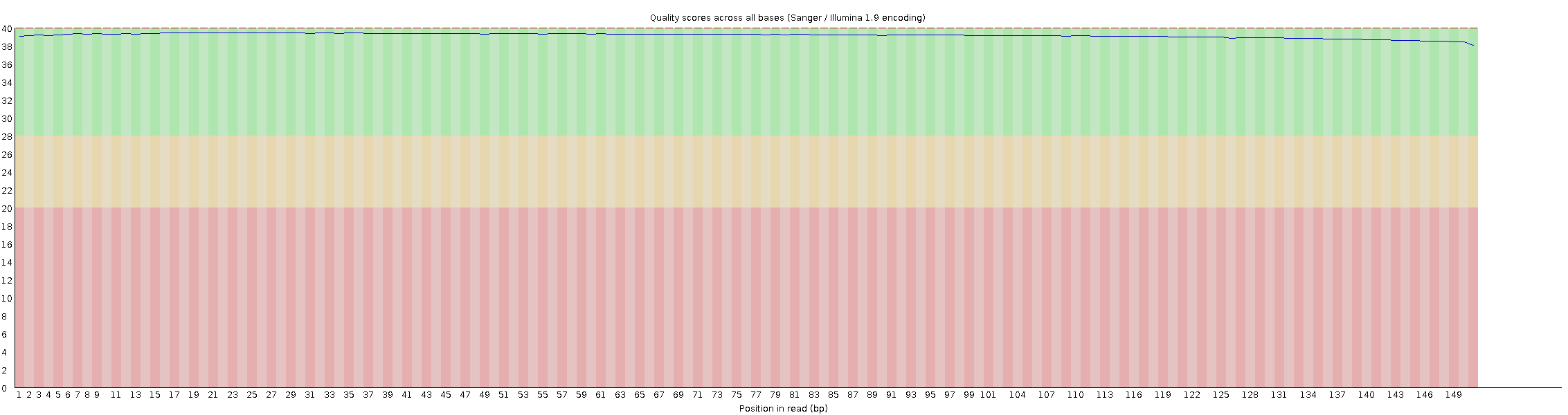

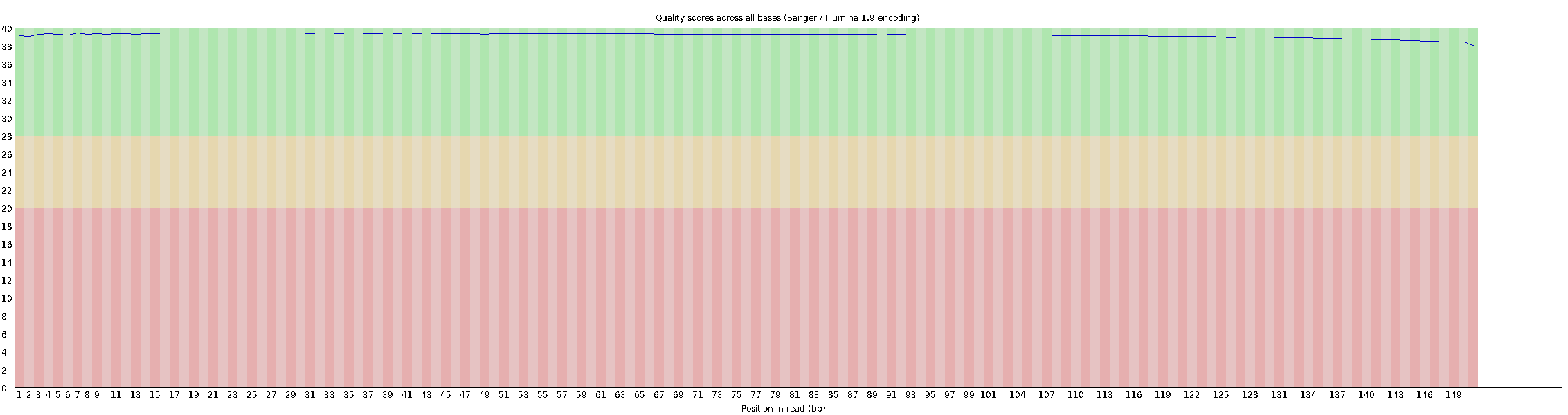

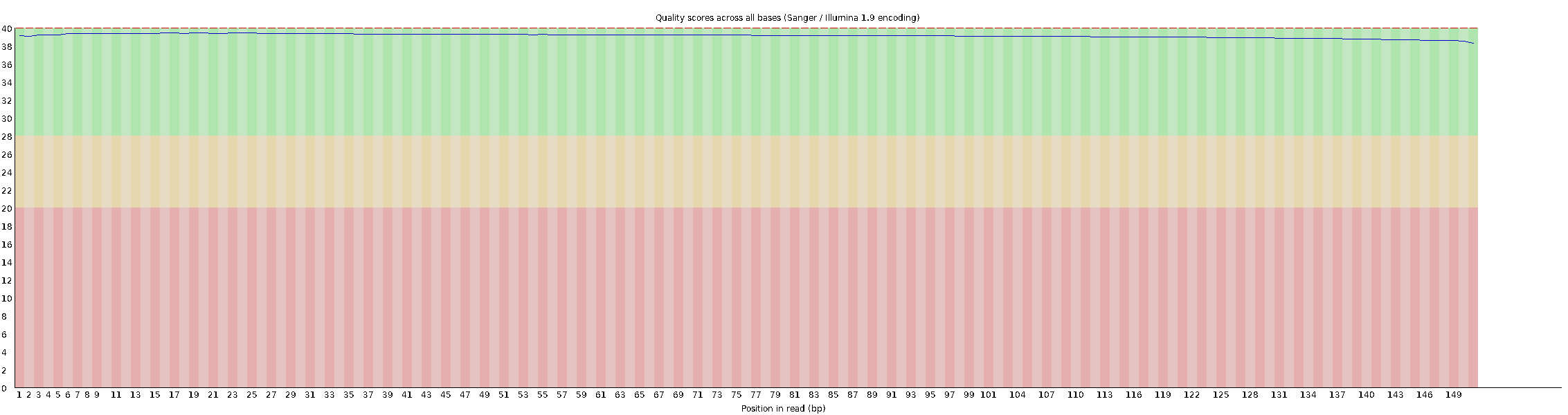

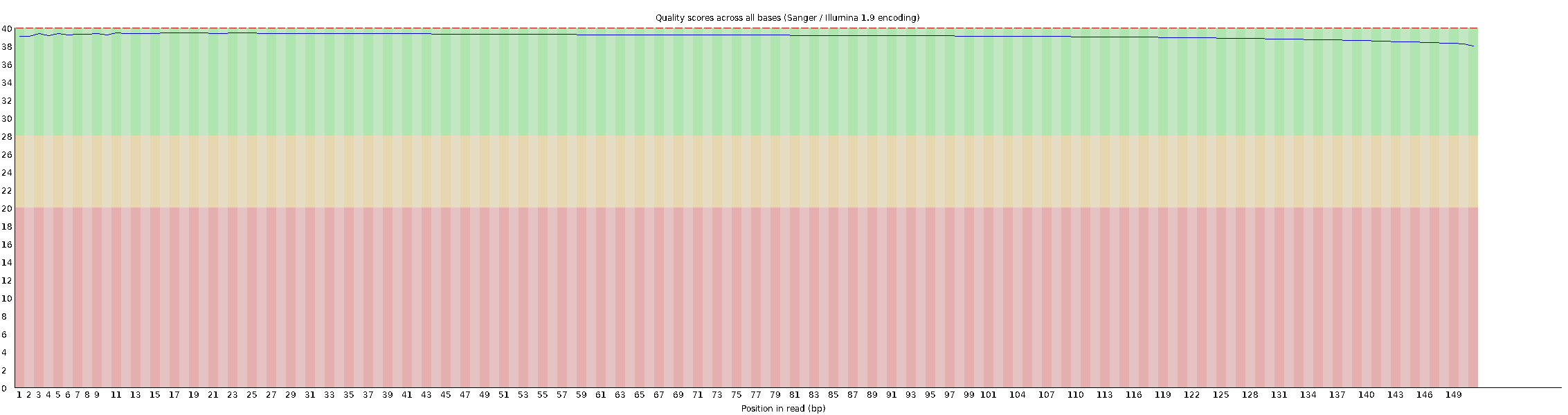

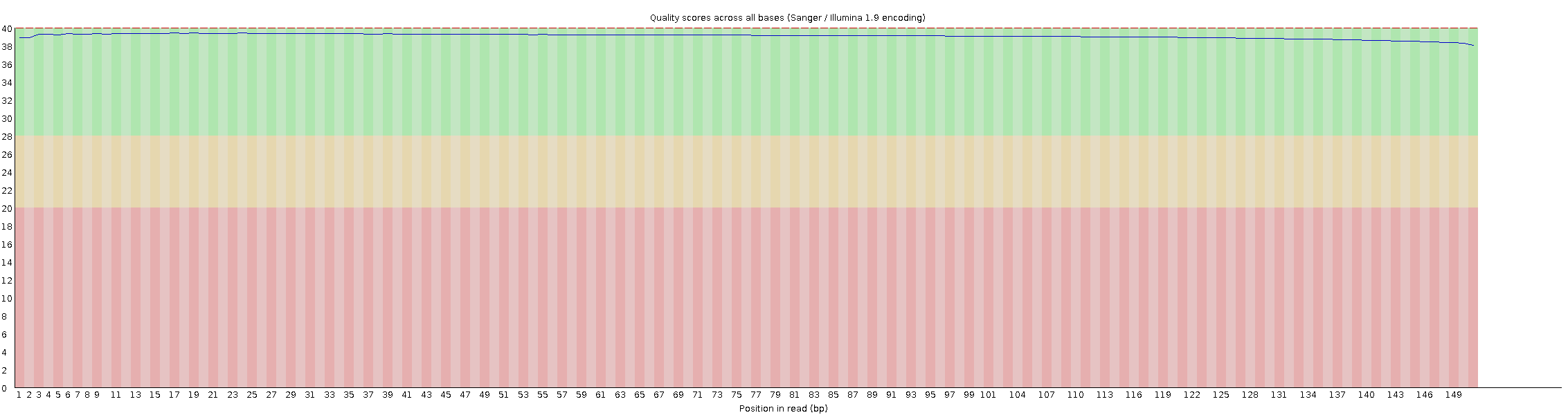

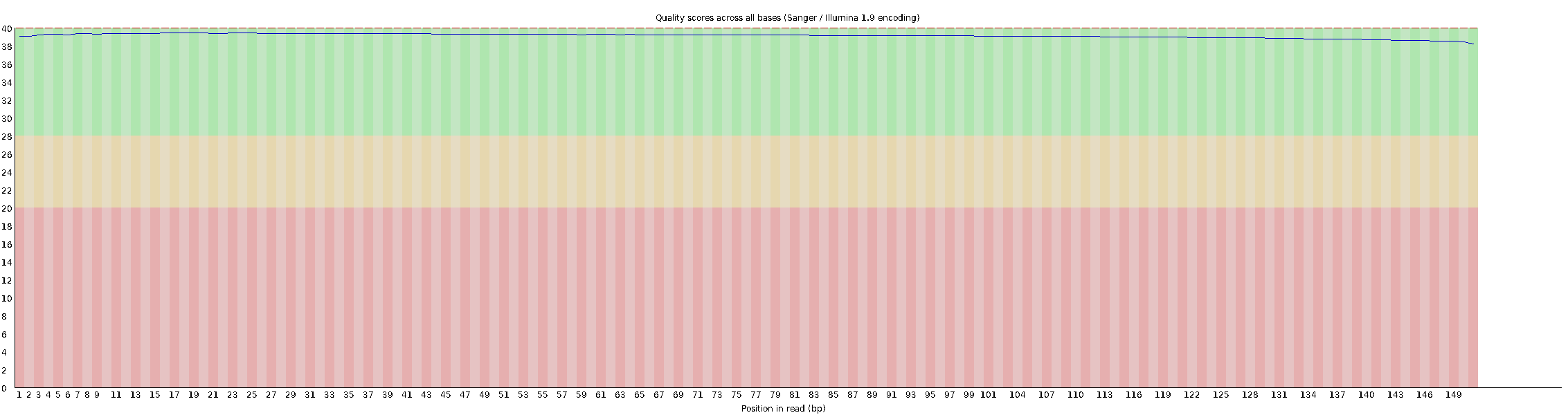

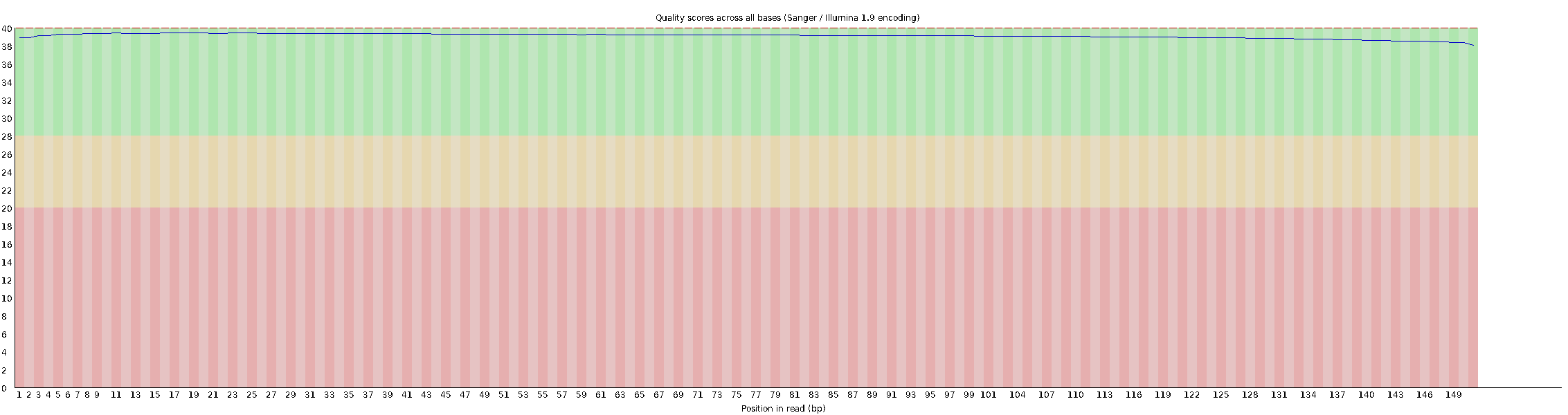

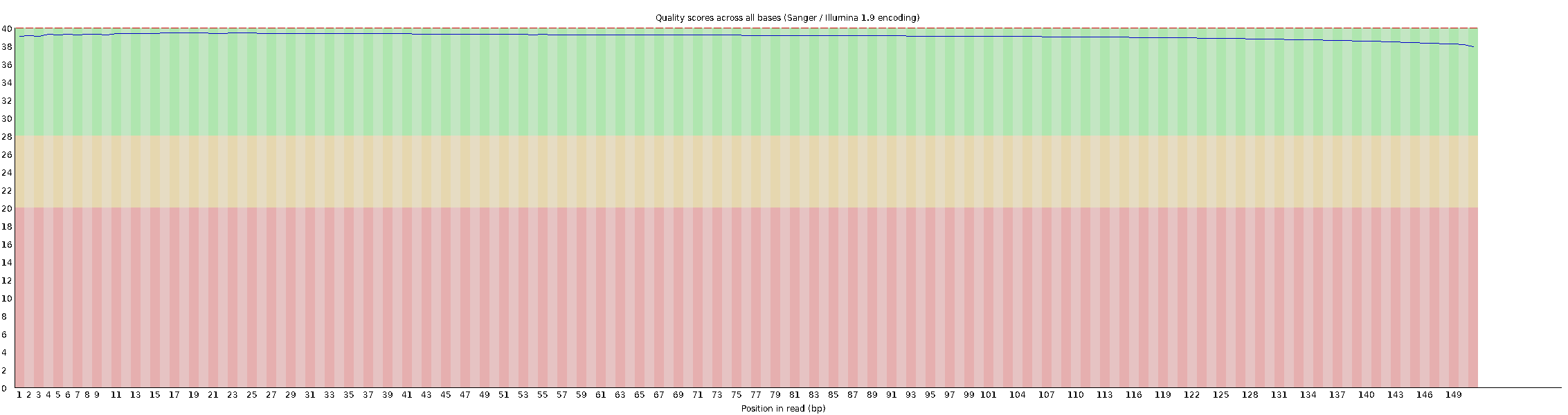

HC1

HC2

HC3

HC3

HC5

L1

L2

L3

L4

L5

P1

P2

P3

P4

P5

**S3: Ethical Approval**

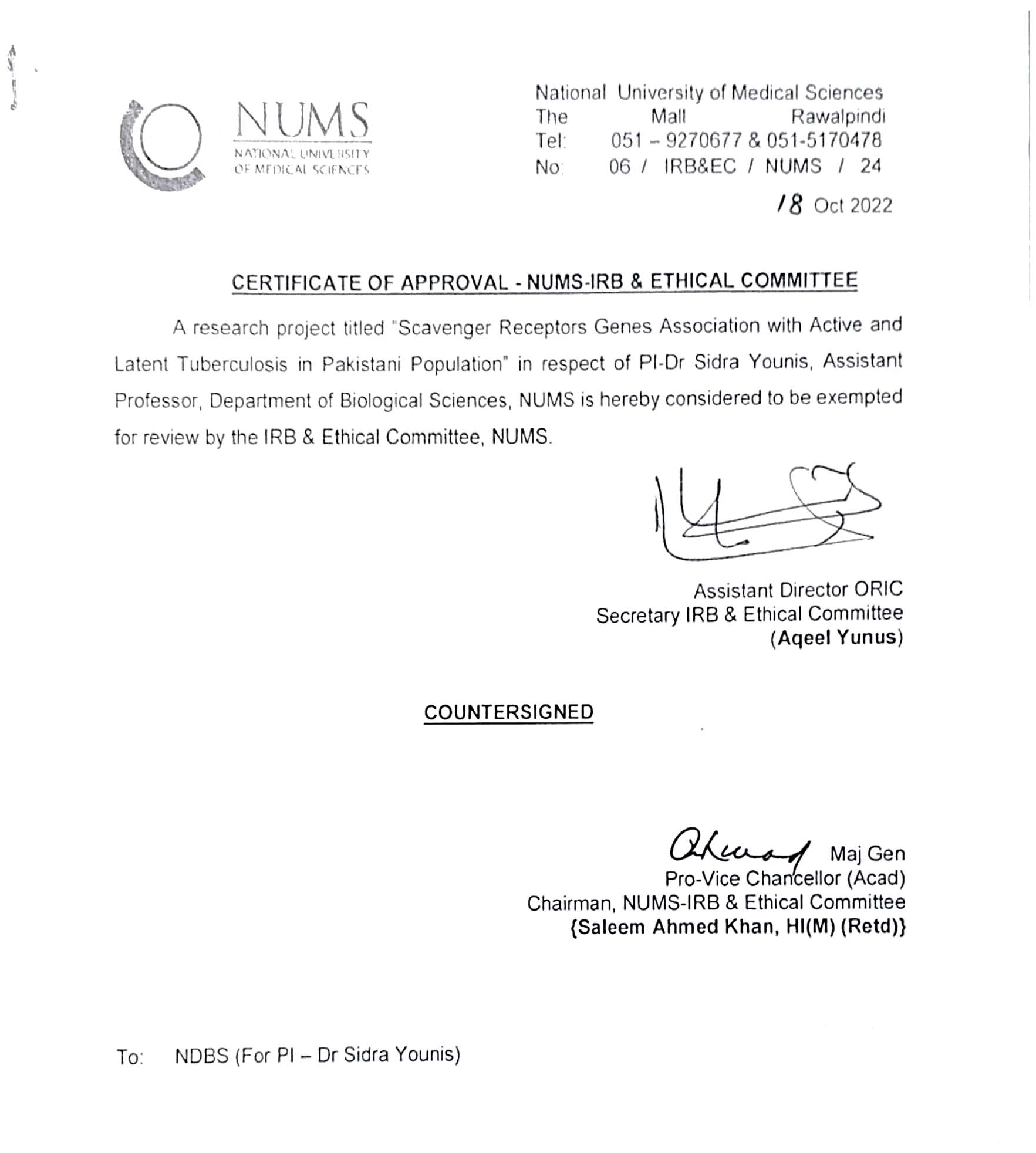

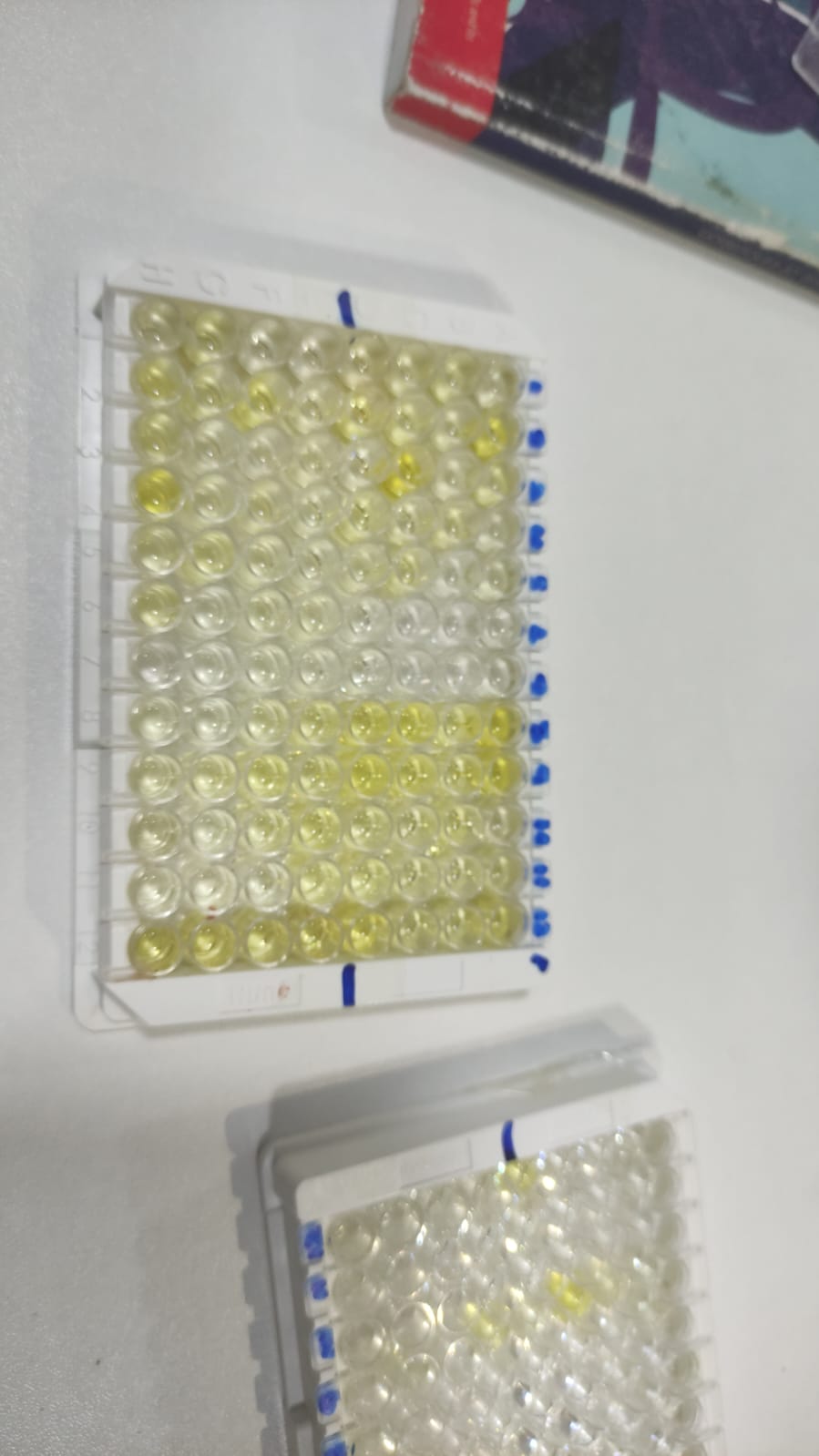

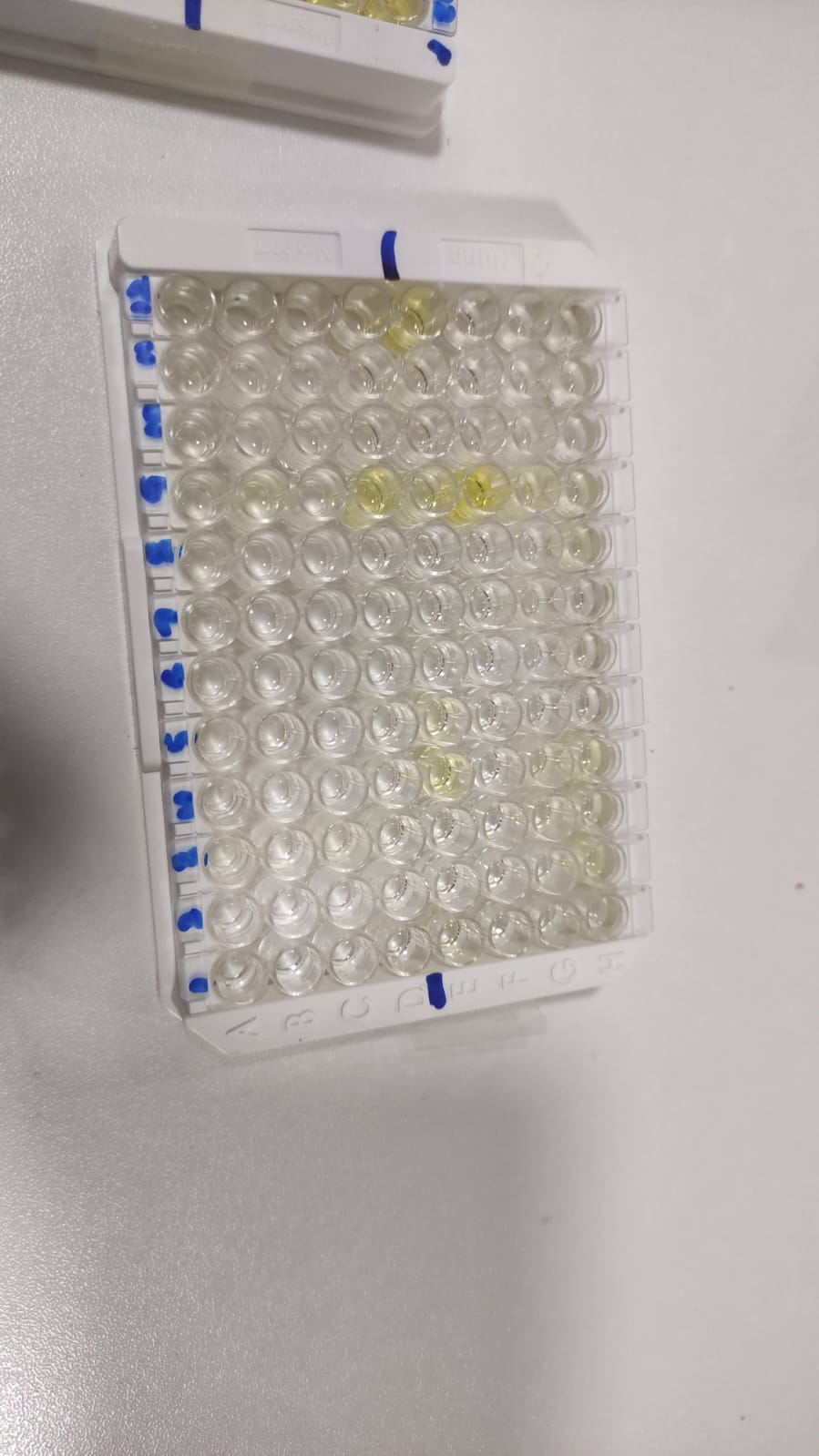
**S4: QUANTIFERON TB Gold Plus Assay**

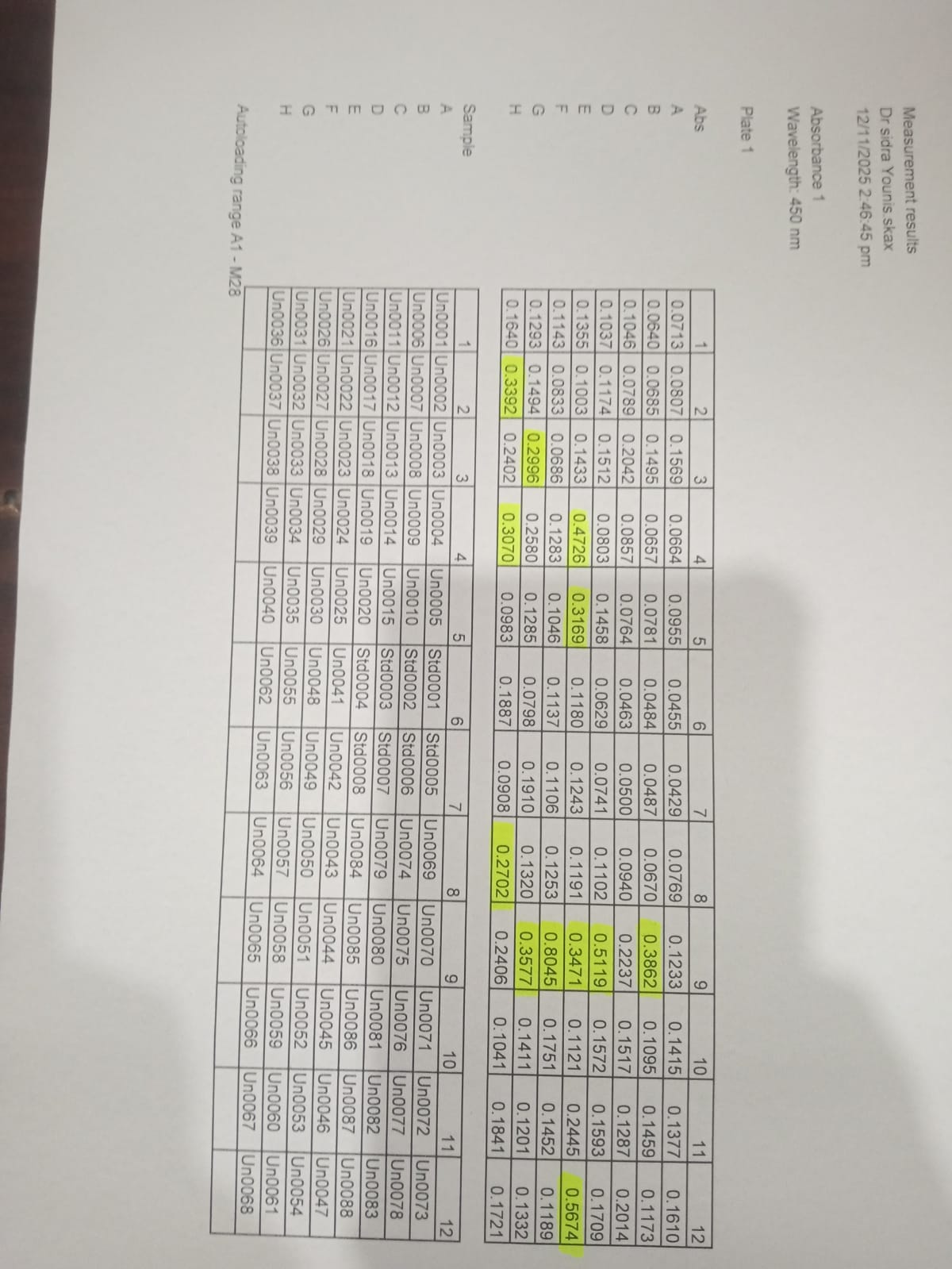

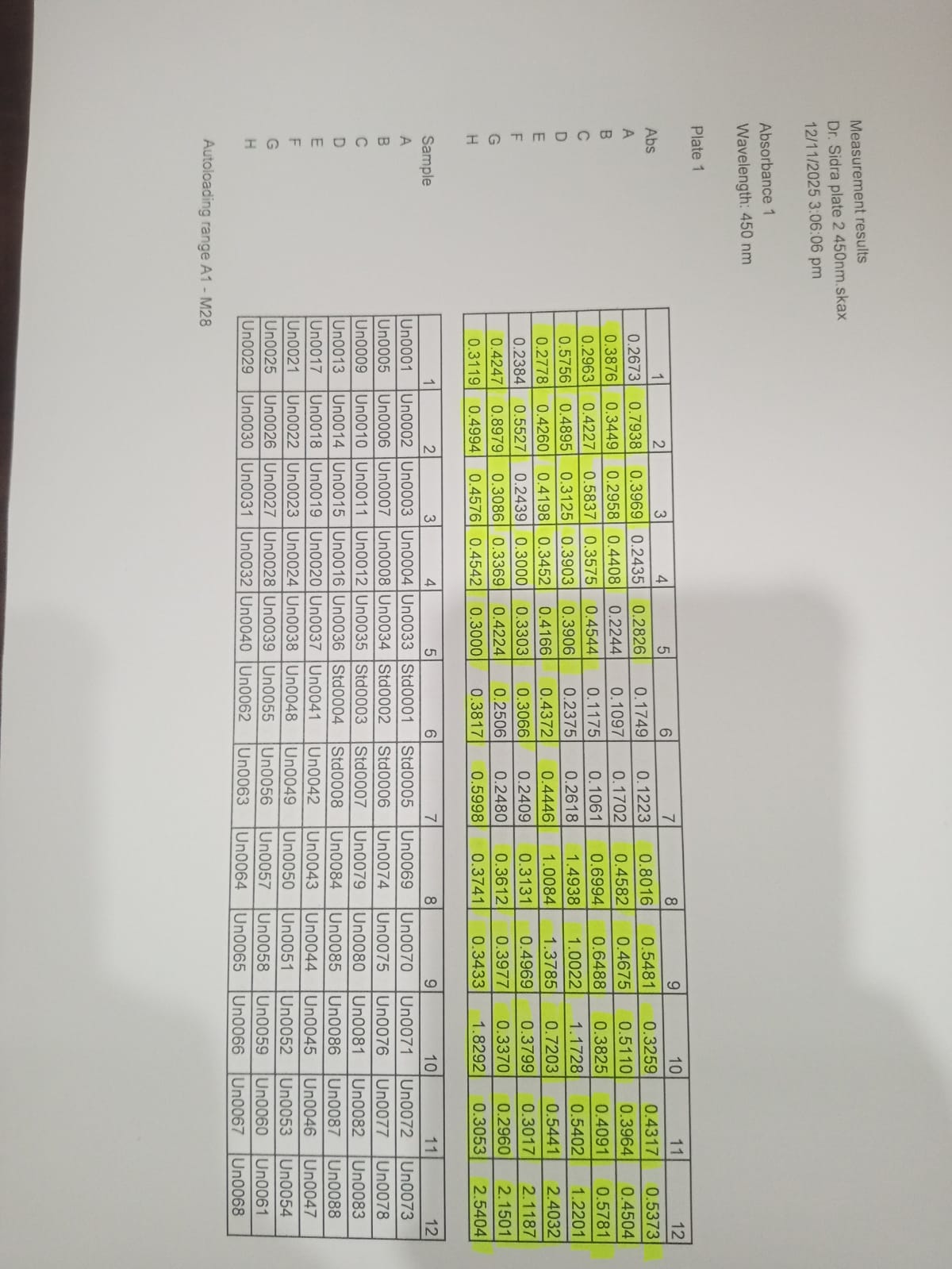
